# Disrupted network segregation of the default mode network in autism spectrum disorder

**DOI:** 10.1101/2021.10.18.21265178

**Authors:** Bo Yang, Min Wang, Weiran Zhou, Xiuqin Wang, Shuaiyu Chen, Lixia Yuan, Marc N. Potenza, Guang-Heng Dong

**Affiliations:** Center for Cognition and Brain Disorders, School of Clinical Medicine and the Affiliated Hospital of Hangzhou Normal University, Hangzhou, Zhejiang Province, PR China; Zhejiang Key Laboratory for Research in Assessment of Cognitive Impairments, Hangzhou, Zhejiang Province, PR China; Department of Psychology, School of Humanities and Social Science, University of Science and Technology of China, Hefei, China; Department of Psychiatry and the Child Study Center, Yale University School of Medicine, New Haven, CT USA; Department of Neuroscience, Yale University, New Haven, CT USA; Connecticut Council on Problem Gambling, Wethersfield, CT USA

**Author notes:** Co-first authors (Bo Yang, Min Wang, Weiran Zhou, Xiuqin Wang).

**Keywords:** Autism spectrum disorders, Default mode network, Participation coefficient

## Abstract

**Background:** Changes in the brain’s default mode network (DMN) in the resting state are closely related to autism spectrum disorder (ASD). Module segmentation can effectively elucidate the neural mechanism of ASD and explore the intra- and internetwork connections by means of the participation coefficient (PC).

**Methods:** Resting-state fMRI data from 269 ASD patients and 340 healthy controls (HCs) were used in the current study. The PC of brain network modules was calculated and compared between ASD subjects and HCs. In addition, we further explored the features according to different age groups and different subtypes of ASD. Intra- and internetwork differences were further calculated to find the potential mechanism underlying the results.

**Results:** ASD subjects showed significantly higher PC of the DMN than HC subjects. This difference was caused by lower intramodule connections within the DMN and higher internetwork connections between the DMN and networks. When the subjects were split into age groups, the results were verified in the 7-12 and 12-18 age groups but not in the adult group (18-25). When the subjects were divided according to different subtypes of ASD, the results were also observed in the classic autism and pervasive developmental disorder groups, but not in the Asperger disorder group. In addition, compared with the HC group, the ASD group showed significantly increased intranetwork connections between the DMN and the frontoparietal network.

**Conclusions:** Less developed network segregation in the DMN could be a valid biomarker for ASD, and this feature was validated with different measures. The current results provide new insights into the neural underpinnings of ASD and provide targets for potential interventions using brain modulation and behavioural training.

## 1. Introduction

Autism is a psychiatric developmental disorder that occurs in early childhood and is characterized by impairments in social communication and interaction, narrow interests and stereotypical repetitive behaviour patterns ^1^. The default network connectivity was enhanced in people with autism when compared to the HC group, which suggested that this enhanced connectivity was associated with the lack of verbal and nonverbal communication ^2,3^. For example, lower strength of posterior cingulate cortex (PCC)-medial prefrontal cortex (mPFC) connectivity in the ASD was associated with poorer social functioning ^4^. Another study found that the strength of intramodule connectivity was significantly lower in the default mode network (DMN) and revealed a high correlation between language and DMN systems ^5^.

The DMN is a large-scale network system in which regions interact instantaneously with sensory, motor and emotional systems to represent the content of imagined events ^6^. Pathological function exists in the DMN of autism spectrum disorder (ASD), involving the mPFC and PCC ^7^. DMN functional pathology is a factor in social cognitive impairment in ASD. Available evidence has revealed abnormalities in the DMN of people with ASD. For example, a study using independent components analysis defined three components and compared the strength of each component/sub-DMN network between groups ^7^. Another study found a voxelwise correlation between PCC and mPFC seeds and the whole brain, which showed decreased FC for the mPFC/anterior cingulate cortex (ACC) in the ASD group ^8^.

Several studies have observed abnormal DMN features at different ASD ages. For example, it has been shown that ASD exhibits hyperconnectivity within the DMN network in primary school children (7-12) and adolescents (12-18) ^9-12^. However, in adults, it has been shown that ASD exhibits hypoconnectivity within the DMN network (19-36) ^12^. A study observed significant differences in internetwork connectivity in children and adolescents with ASD compared to adults ^13^. In terms of development as a whole, there is evidence in the literature of a nonlinear downward trend (rising, then falling); specifically, the modularity of ASD rises from children to adolescents and then falls from adolescents to adults ^14^.

According to the DSM-IV, ASD includes 3 subtypes: 1) autistic disorder (classic autism), 2) Asperger’s disorder (language development at the expected age, no mental retardation), 3) pervasive developmental disorder (PDD), and not otherwise specified (individuals who have autistic features and do not fit any of the other subtypes) ^15^. A study found that high-functioning autism (HFA) and Asperger’s syndrome (AS) are characterized by complex, age-related developmental differences, mainly in quantitative and qualitative differences in language development and cognitive functioning ^16^. When mapping network dynamics onto time series in the behavioural domain, abnormalities were found in the emotional and visual behavioural subdomains, which were more pronounced in people with autism compared to Asperger’s syndrome within the ASD spectrum ^17^. At the same time, there are studies proving that PDD has a high degree of variability in clinical presentation ^18^. Subgroups are complex, with inconsistent phenotypes between subgroups, large individual differences, and even greater variability in structural and functional aspects of the central nervous system. Therefore, we need to perform further analysis of the neural mechanisms of each subtype to identify differences.

The participation coefficient (PC) is a summary network topology measure that quantifies how connected a defined ROI is to other ROI inter- and intranetworks. The PC allows us to react more visually and clearly to the connections between networks. An internetwork is a cross-network in which more connections are made, which indicates network integration. An intranetwork exists when there are more connections within the network, which indicates network segregation. Internetwork anomalies have been widely verified in other psychiatric disorders (e.g., major depressive disorder, anxiety disorders and schizophrenia), showing decreased PC in the DMN and reflecting poor modular segregation between resting-state networks ^19-21^. Most psychiatric disorder studies tend to use this kind of large-scale network to avoid single-seed analysis distortion of the interpretation of the whole network. Therefore, PC analysis of the development of passive resting brain networks may provide a comprehensive perspective for the study of ASD.

Most of the studies have only been conducted on one (or two) subtypes or age groups, which does not provide systematic knowledge for understanding ASD. In the current study, we first analysed the intra- and internetwork differences in all subjects; then, we explored the features in different age groups and in different subtypes of ASD. The following assumptions were made: 1) in all subjects, there may be a significant change in the DMN in the ASD group; 2) as age increases, the modularity of the DMN in ASD may increase from childhood to adolescence and then fall from adolescence to adulthood; and 3) there may be differences among the subtypes of ASD.

## 2. Methods

### 2.1. Participants

The dataset in this study originated from the initial Autism Brain Imaging Data Exchange (ABIDE I). ABIDE I involved 24 international sites with 1,112 subjects, including 539 ASD patients and 573 healthy controls (HCs) (ages 7-64 years, median 14.7 years across groups). Functional and structural brain imaging datasets and phenotypical datasets were downloaded from the ABIDE website (http://fcon_1000.projects.nitrc.org/indi/abide/).

The exclusion criteria for subjects were as follows: 1) without functional or structural images; 2) without handedness information or with mixed handedness; 3) without full-scale intelligence quotient (FIQ) information or FIQ smaller than 70 ^22^; 4) eye status, time of repetition (TR), slice number, or data matrix size different from those of most subjects within a site; we used 180 time points for all Stanford subjects; 5) time points different from those of most subjects within a site (at the Stanford site, there were 20 subjects with 240 time points (50%), 17 subjects had 180 time points (42.5%), one subject had 181 time points, two subjects had 238 time points, and we truncated all the functional data to 180 time points.); 6) severe artefacts and signal losses in functional images (by visual inspection); 7) scan duration less than 100 time points ^23^; 8) head motion exceeding 3 mm or 3 degrees; 9) bad spatial normalization (by visual inspection); 10) scan cover less than 91% of the whole brain; 11) spatial correlation < 0.6 (a threshold defined by mean - 2SD) between each participant’s regional homogeneity (ReHo) map and the group mean ReHo map ^24^; 12) some subjects were excluded to ensure group matching for age, FIQ, mean framewise displacement (mFD)^25^ (*p* > 0.05, Two-sample t-test) and sex and handedness (*p* > 0.05, Chi-square test) in each centre; 13) at each step, any sites (UCLA_2 after poor spatial normalization, Caltech after inadequate cover) with less than 20 individual datasets were excluded; 14) subjects were screened according to DCM-IV (sites with a DSM-IV score of -9999 and a certain invalid score were excluded) and age (7-25 years); ultimately, a total of 609 subjects, including 269 ASDs and 340 HCs at 13 sites, were included in our investigation. The reserved subjects after each step can be found in Fig. 1.

**Fig. 1.**
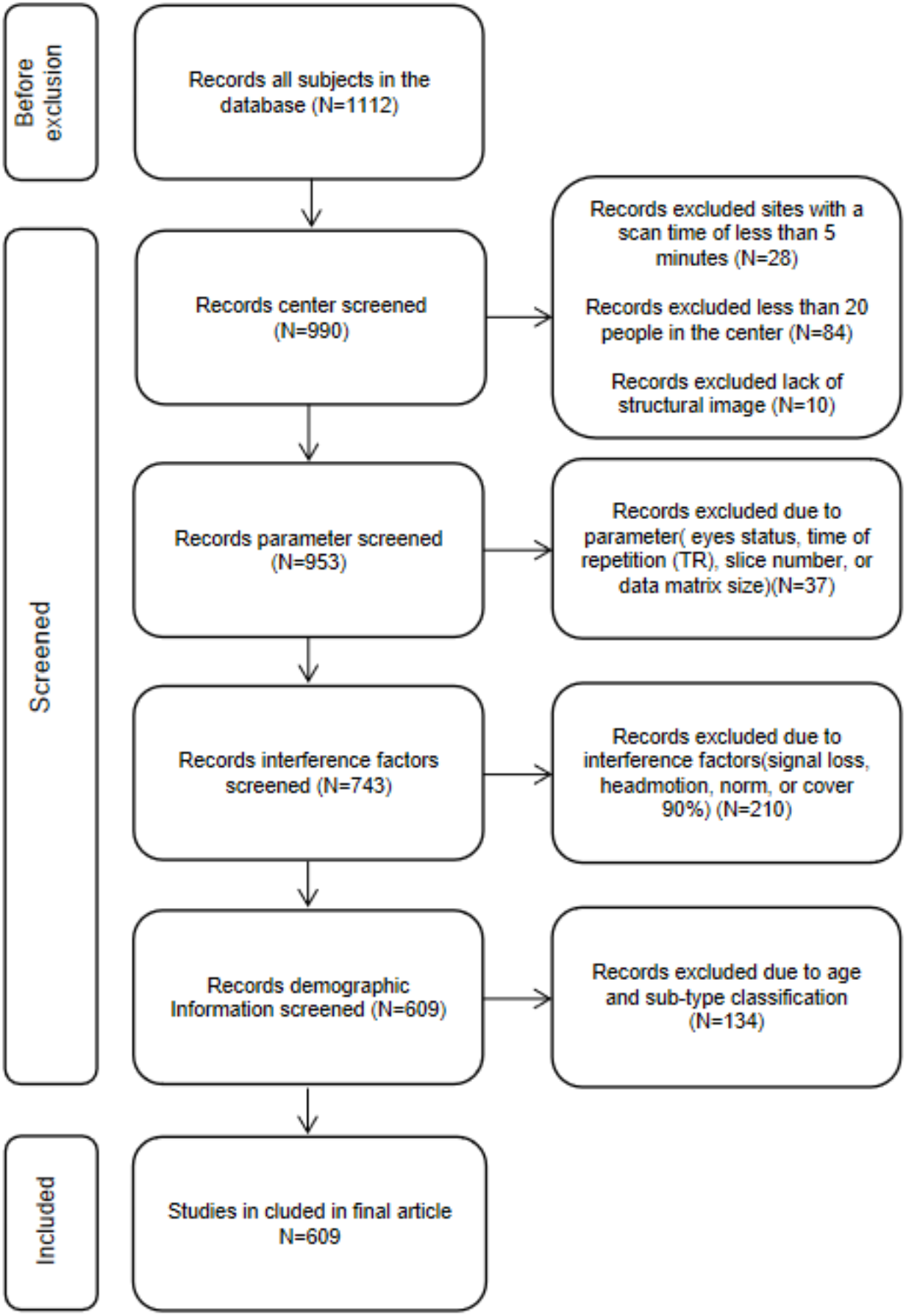
Screening flowchart of the subject selection in current study. The subplot shows that criteria for the screening of 1112 subjects downloaded from the ABIDE webpage.

### 2.2. MRI Data Preprocessing

Data preprocessing was performed with DPABI V5.1 (http://rfmri.org/dpabi) and SPM12 (http://www.fil.ion.ucl.ac.uk/spm/software/spm12/). Preprocessing steps included: 1) discarding the first 10 time points; 2) slice timing correction; 3) head motion correction; 4) spatial normalization with the forward transformation field from the unified segmentation of anatomic images, and then resampling into 3 × 3 × 3 mm^3^); 5) spatial smoothing with a 3D isotropic Gaussian kernel with a full width half max (FWHM) of 6 mm; 6) removing the linear trend; 7) nuisance covariates regression, including head motion covariates with Friston 24-parameter model and white matter and cerebrospinal fluid signal; 8) filtering the data with a passband filter of 0.01-0.08 Hz.

### 2.3. Network construction and graph theory analysis

Based on the network construction method in a previous study ^26^, we applied a functional template ^27^ to obtain defined nodes. This functional template allowed the brain to be divided into 160 functionally isolated regions of interest (ROIs) covering a large portion of the cerebral cortex and cerebellum. The set of ROIs (3 mm diameter spheres) was generated using the peak coordinates obtained from a meta-analysis of a series of fMRI activation studies ^27^. Whole-brain networks were computed for each participant using CONN (https://www.nitrc.org/projects/conn/). The BOLD signal was extracted from each ROI, and the bivariate correlation between each pair of ROIs was calculated to obtain a 160×160 correlation matrix for each participant (see Fig. 2B). To be able to compare network properties between participants and groups, we chose a 15% sparsity threshold, which was favoured by previous researchers. Such a threshold allowed for a balance between using very sparse graphs and denser graphs ^28^.

**Fig. 2.**
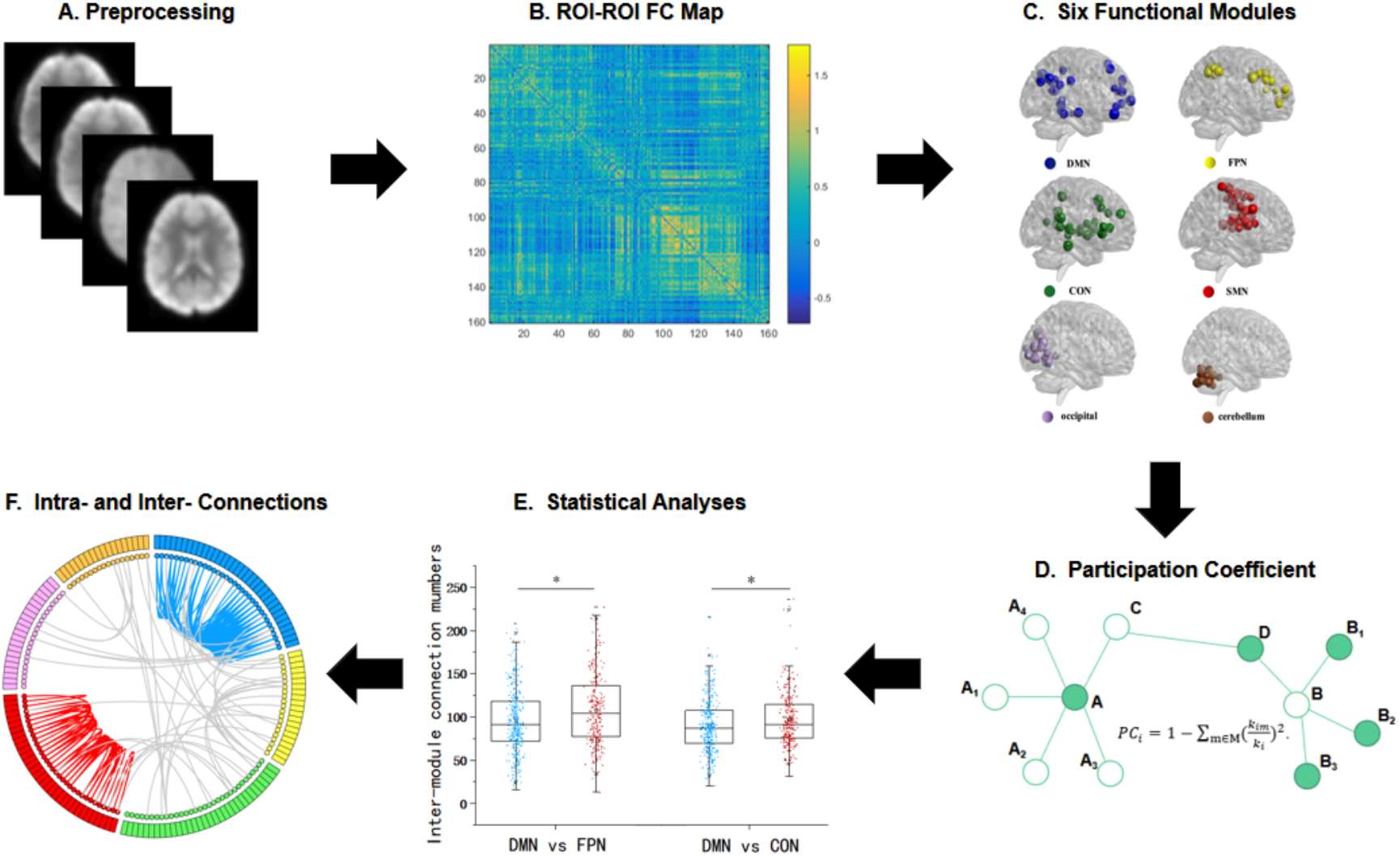
The experimental flowchart in current data analyses. The subplot (A) shows the process of obtaining FMRI images. The subplot (B) shows the application of the Dosenbach 160 ROI functional template to the defined nodes and the calculation of the task-related ROI-ROI functional connectivity. Subplot (C) shows the module partitions: the default-mode network (DMN), frontal-parietal network (FPN), cingulo-opercular network (CON), sensorimotor network (SMN), occipital network, and cerebellum. The subplot (D) shows the calculation and comparison of the participation coefficient (PC) between the autism spectrum disorder (ASD) group and the healthy control (HC) group. Subplot (E) shows the statistical plots for analysing significant differences in PC. The subplot (F) shows whether PC differences between the two groups are driven by changes in intramodule connections and intermodule connections

All graph theory measures were calculated using the Gretna Toolbox ^29^. According to a previous study ^27^, 160 ROIs were assigned to six functional modules corresponding to the default-mode network (DMN), frontoparietal network (FPN), cingulate-opercular network (CON), sensorimotor network (SMN), occipital network (OCC) and cerebellum (CER). We used this modular structure to compute the following graph theory measures. First, the participation coefficient (PC) was calculated to quantify the degree of modular segregation. For node i, PC is defined as 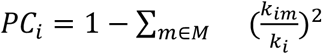 (see Fig. 2D). In this formula, m is a module in a set of modules M, k_im_ is the number of connections between node i and module m, and k_i_ is the total number of connections of node i in the entire brain network ^30^. PC_i_ measures the proportion of inter- and intramodule connections for node i. In addition to the PC, we also counted the number of connections within each module, as well as the number of connections between any two pairs of modules. In general, if a node is highly integrated with nodes in its own module but less integrated with nodes in other modules (higher degree of modular isolation), the PC is close to zero; conversely, if a node is less integrated with nodes in its own module but more integrated with nodes in other modules (lower degree of modular isolation), the PC will be close to 1. Here, we use each module’s average PC to characterize modular segregation.

### 2.4. Multisite effect correction

To account for site, collection time, and data acquisition parameter variability across each of the data collections in ABIDE I, site effects on PC were removed using the ComBat function available in MATLAB ^31^ (https://github.com/Jfortin1/ComBatHarmonization). This approach has been shown to effectively account for scanner-related variance in multisite resting-state fMRI datasets ^31^. During Combat, the diagnosis, age, sex, FIQ, and mFD were treated as biological variables of interest, and a default nonparametric prior method was used in the empirical Bayes procedure.

### 2.5. Classification

According to the present law of age development, we divide age into three groups: primary school age of 7-12 years, adolescents aged 12-18 years, and young adults aged 18-25 years.

The 3 DSM-IV ASD subtypes are 1) autistic disorder (classic autism), 2) Asperger’s disorder, and 3) pervasive developmental disorder (PDD) ^15^. Our study divided the ASD group into three subtypes based on DSM-4 (type I, type II, and type III as corresponding abbreviations).

### 2.6. Statistical analyses

First, for between-group comparisons of demographic information and mean PCs, two-sample t-tests were conducted using SPSS Version 22.0 (IBM Corporation, Armonk, NY, USA). We then used two-sample t-tests to analyse changes in the number of intramodule connections and intermodule connections for modules that differed significantly between the two groups. To control for multiple comparisons, a Bonferroni correction was used, and the significance threshold was set at α = 0.05/6 (6 measurements) = 0.0083.

Second, to explore the relationship between different age groups (or subtypes) and HC, the correlation between the mean PC in subjects was compared (a significant outcome for comparison between the two groups).

Finally, to more specifically explain the reasons for PC changes, building on the previous analysis, we further used two-sample t-tests to analyse the number of inter- and intramodule connections.

## 3. Results

### 3.1. Module partitions

As shown in Fig. 2C, the 160 ROIs were divided into six functional modules, including the default mode network (DMN), frontoparietal network (FPN), cingulo-opercular network (CON), sensorimotor network (SMN), occipital network (OCC) and cerebellum (CER) ^32^.

### 3.2. PC results between ASD and healthy controls

Overall, we compared the mean PC in the HC and ASD groups. Specifically, the mean PC of the DMN was significantly higher in the ASD group than in the HC group during passive resting conditions (DMN: *t* = -5.352, *p* _corrected_ = 0.000) (see Fig. 3A). There was no difference in the PC between HC and ASD in the other modules.

**Fig. 3.**
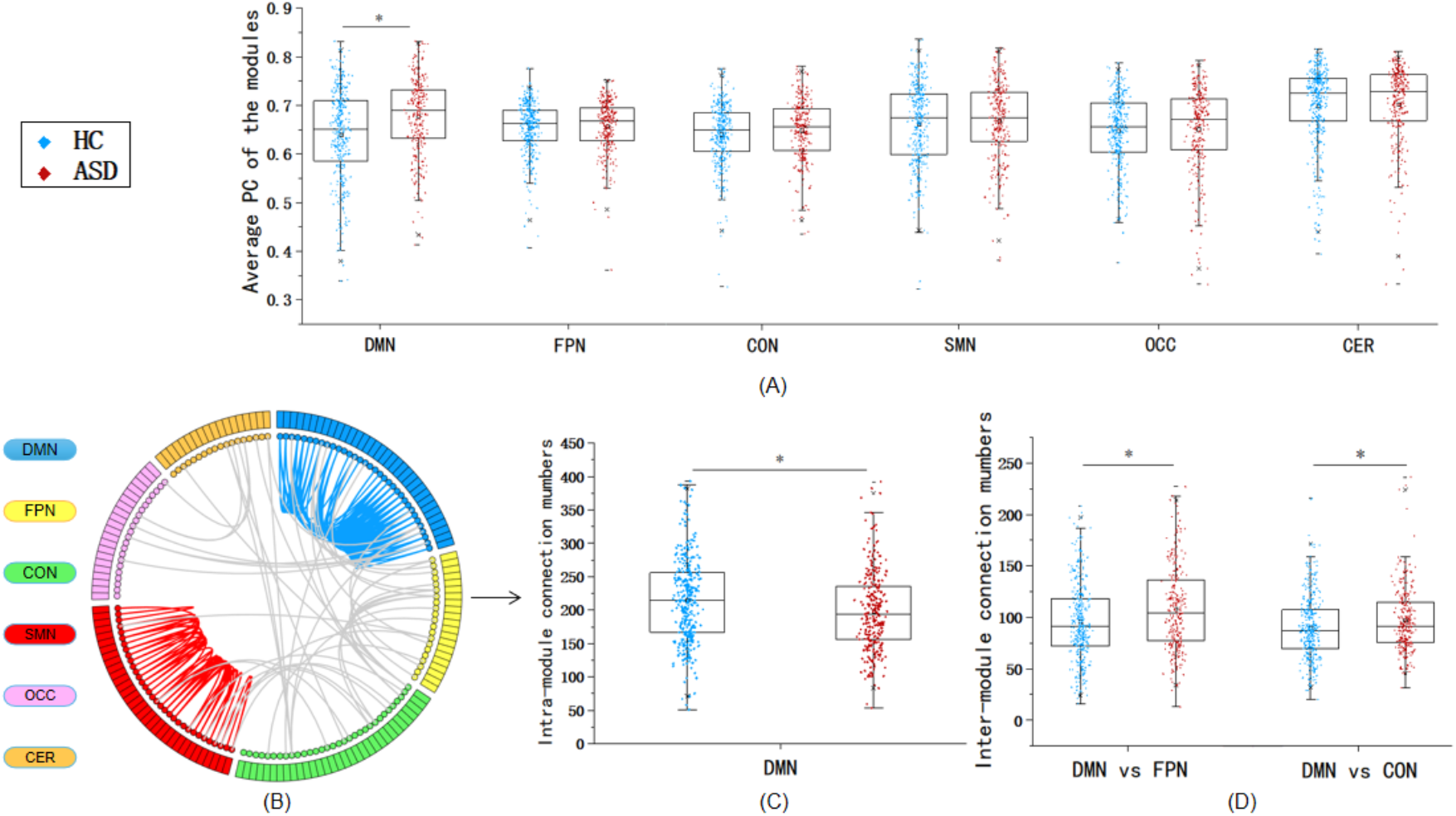
Mean participation coefficients and intramodal and intermodal connections differed between the HC and ASD groups in all subjects. The subplot (A) shows that during passive resting conditions, compared with the healthy control (HC) group, the autism spectrum disorder (ASD) group showed a significantly increased participation coefficient (PC) on the DMN. The subplot (B) shows an example of the intermodular and intramodular connections of the default-mode network (DMN) and sensorimotor network (SMN). Subplot (C) shows that from an intramodular perspective, there were fewer connections within the DMN network and SMN network in the ASD group than in the control group. The subplot (D) shows that from an intermodular perspective, compared with the HC group, the autism spectrum disorder (ASD) group showed a significantly increased DMN with FPN (or CON). * Indicates results can survive Bonferroni correction (*P<*0.008).

Furthermore, we examined whether this difference between the two groups was driven by variations in intramodule connections, intermodule connections or a combination of both. As shown in Fig. 3C, the ASD group had significantly fewer connections within the DMN module than the HC group (*t*=3.372, *p* _corrected_=0.001). However, specificity was observed in intranetwork connections of the SMN (*t*=3.021, *p* _corrected_=0.003) (Fig. 3C). In the other modules, there was no difference in the intranetwork connection between HC and ASD.

Compared with the HC group, the autism spectrum disorder (ASD) group showed a significantly increased DMN with FPN (or CON) (*t*=-3.988, *p* _corrected_=0.000, *t*=-2.742, *p* _corrected_=0.006) (Fig. 3D) interconnections. There was no difference in the internetwork connection between HC and ASD in the other two-by-two modules.

### 3.3. The PC features in different age groups

The ASD cohort was stratified into three age groups (primary school aged 7-12 years, adolescents 12-18 years, young adults 18-25 years). We compared the mean PC in the HC and ASD groups in each age group separately. The mean PC of the DMN was significantly higher in the ASD group than in the HC group in primary school-aged children (DMN: *t* = -2.689, *p* _corrected_ = 0.008) and adolescents (DMN: *t* = -4.284, *p* _corrected_ = 0.000) (see Fig. 4A). There were no significant differences among young adults for all modules, including the DMN.

**Fig. 4.**
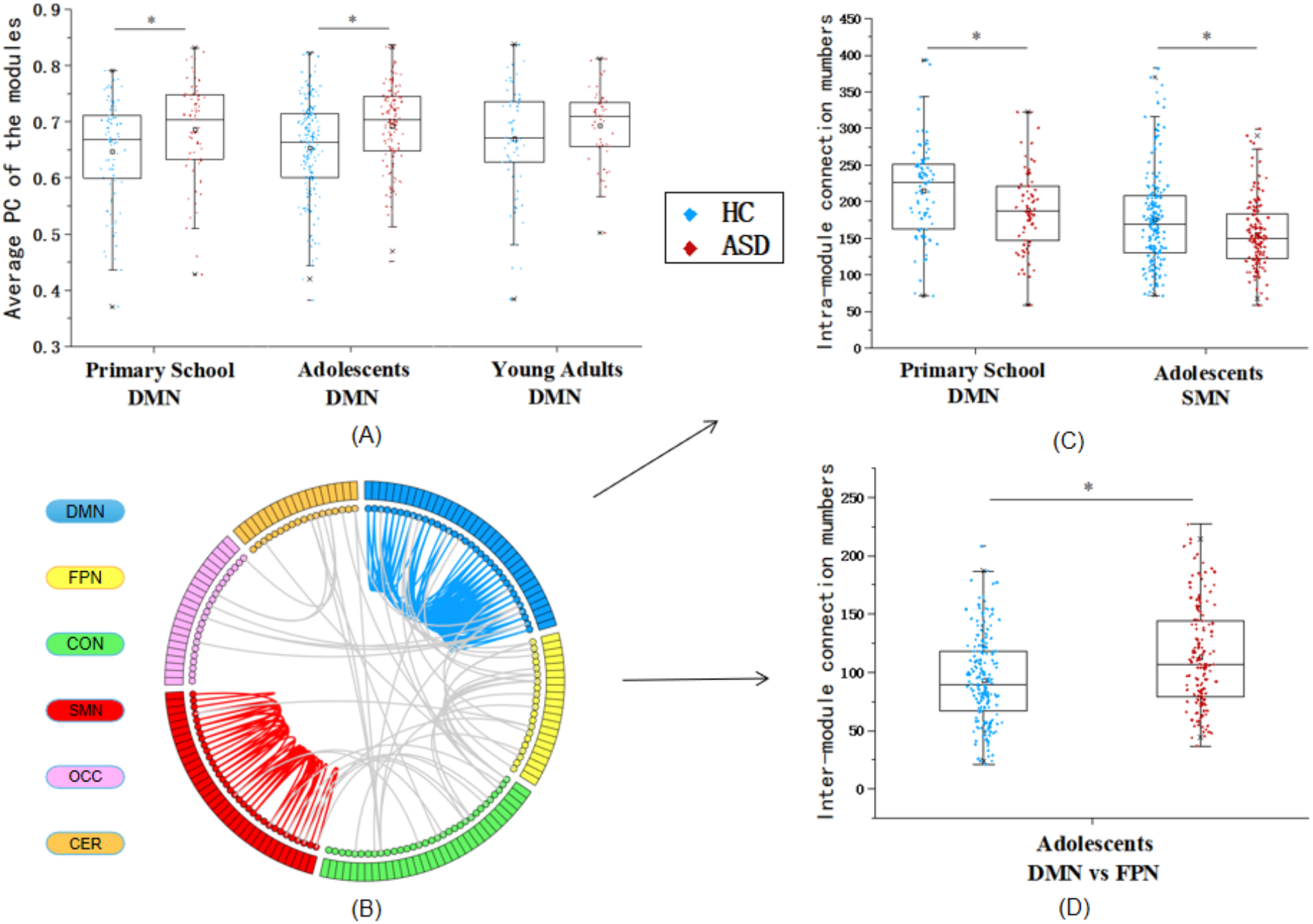
Mean participation coefficients and intramodal and intermodal connections between the HC and ASD in different age groups. Subplot (A) shows a higher mean PC of the DMN in the ASD group than in the HC group in primary school-aged adolescents. Subplot (B) shows an example of the intermodular and intramodular connections of the default-mode network (DMN) and sensorimotor network (SMN). Subplot (C) shows that from an intramodular perspective, compared to the HC group, there were fewer connections within the DMN network in the primary school-aged group and the SMN network in adolescents in the ASD group. The subplot (D) shows that from an intermodular perspective, compared with the HC group, the autism spectrum disorder (ASD) group showed significantly increased DMN with FPN in adolescents. * Indicates results can survive Bonferroni correction (*P<*0.008).

Furthermore, for ASD subjects in the primary school age group, we found that the ASD group had significantly fewer connections within the DMN module than the HC group (*t*=2.734, *p* _corrected_=0.007). Specificity was observed in intranetwork connections of SMN in adolescents (*t*=3.314, *p* _corrected_=0.001) (Fig. 4C). Meanwhile, compared with the HC group, the autism spectrum disorder (ASD) group showed significantly increased DMN with FPN in adolescent (*t*=-4.377, *p* _corrected=_0.000) (Fig. 4D) interconnections. There was no difference in the internetwork connection and intermodule connections between HC and ASD in the other age groups.

### 3.4. The PC features in different subtypes of ASD

According to the DSM-5, subjects were classified into three subtypes (Type I, Type II, Type III). The mean PC of the DMN was significantly higher in the ASD group than in the HC group in Type I (DMN: *t* = -4.641, *p* _corrected_ = 0.000) and Type III (DMN: *t* = -3.629, *p* _corrected_ = 0.001) (see Fig. 5A). There were no significant differences in Type II for all modules, including the DMN.

**Fig. 5.**
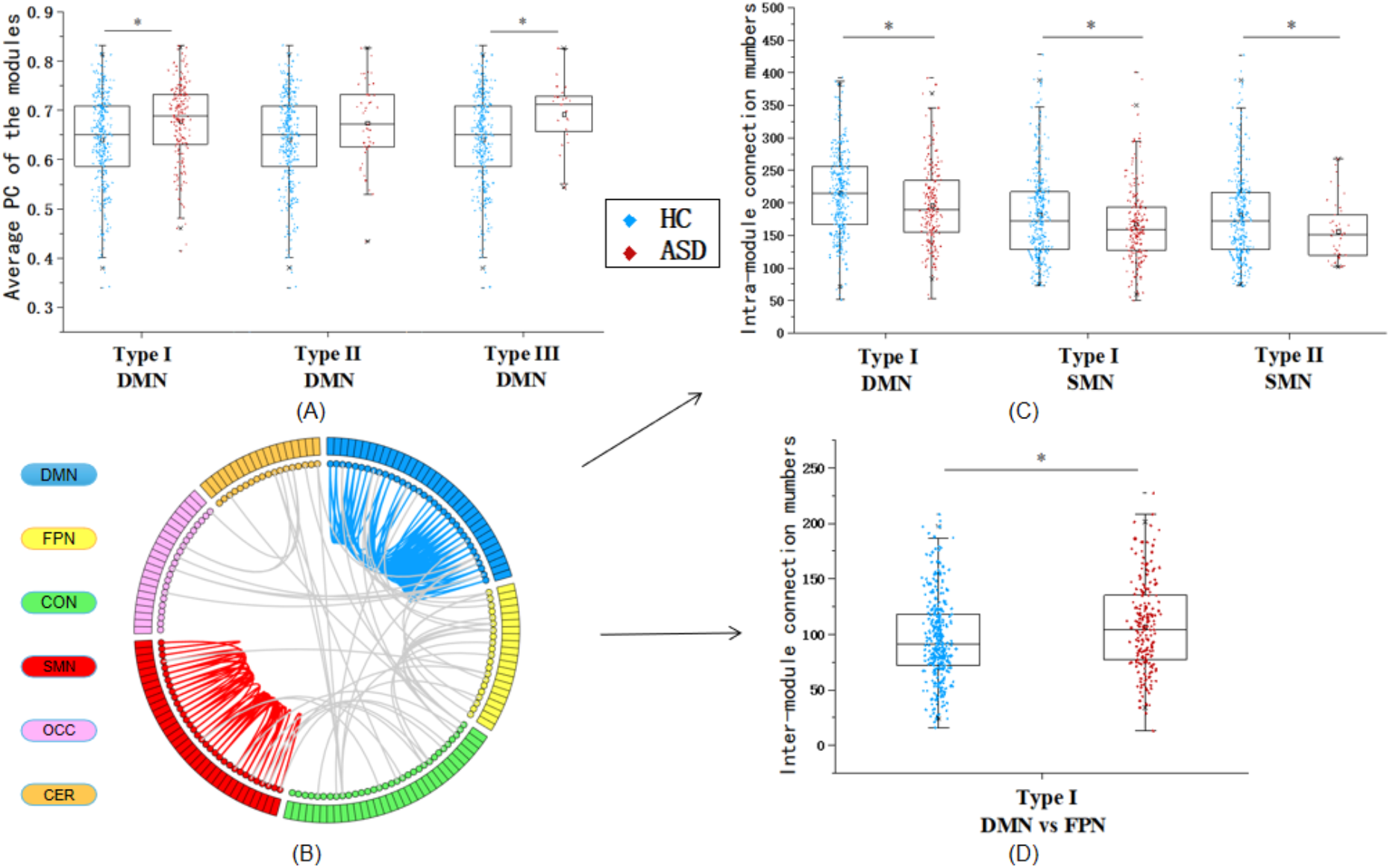
Mean participation coefficients and intramodal and intermodal connections between the HC and different subtype ASD groups. Subplot (A) shows a higher mean PC of the DMN in the ASD group than in the HC group in Type I and Type III. Subplot (B) shows an example of the intermodular and intramodular connections of the default-mode network (DMN) and sensorimotor network (SMN). Subplot (C) shows that from an intramodular perspective, compared to the HC group, there were fewer connections within the DMN network in Type I and the SMN network in Type I (and Type II) in the ASD group. The subplot (D) shows that from an intermodular perspective, compared with the HC group, the autism spectrum disorder (ASD) group showed a significantly increased DMN with FPN in Type I. * Indicates results can survive Bonferroni correction (*P<*0.008).

Furthermore, when ASD subjects were classified into three subtypes (Type I, Type II, Type III), we found that the ASD group had significantly fewer connections within the DMN module than the HC group in Type I (*t*=3.44, *p* _corrected_=0.001) (Fig. 5C). Specificity was observed in intranetwork connections of SMN in Type I and Type II (*t*=2.745, *p* _corrected_=0.006, *t*=3.321, *p* _corrected_=0.001) (Fig. 5C). Compared with the HC group, the autism spectrum disorder (ASD) group showed significantly increased DMN with FPN (*t*=-3.387, *p* _corrected_=0.001) (Fig. 5D) interconnections in Type I. There was no difference in the internetwork connection and intermodule connections between HC and ASD in the other subtype groups.

### 3.5. Replication test with an independent sample

We compared the mean PC in the HC and ASD groups from another database (ABIDE II). Specifically, the mean PC of the DMN was significantly higher in the ASD group than in the HC group during passive resting conditions (DMN: *t* = 2.348, *p* _corrected_ = 0.021), which validated our results (Fig. 6A). Further analysis showed that the ASD group had significantly fewer connections within the SMN module than the HC group (*t*= 2.006, *p* _corrected_= 0.048), which validated some of our results (Fig. 6B). The results replicated the main findings in the current study and proved that the current results from a large sample are solid.

**Fig. 6.**
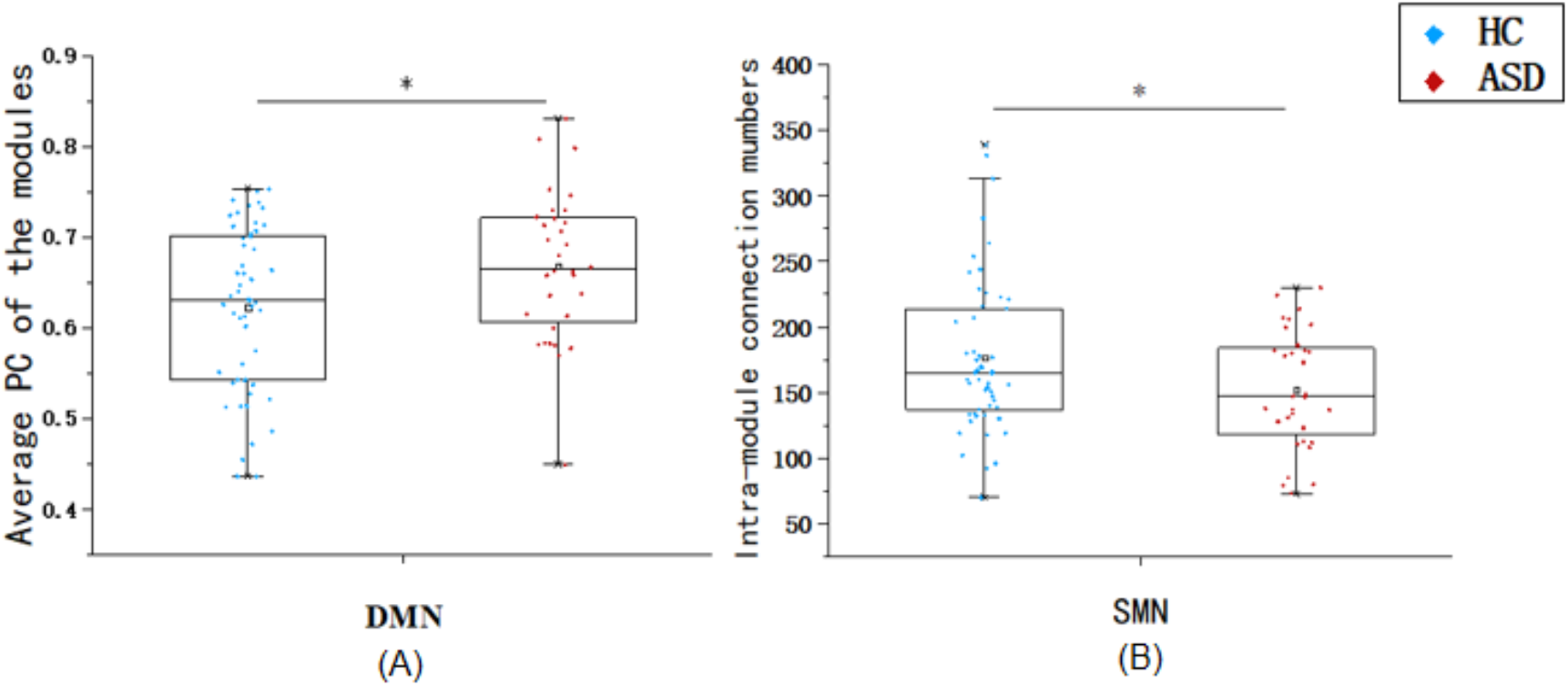
Replication test on mean PC and intranetwork test results. Subplot (A) shows a higher mean PC of the DMN in the ASD group than in the HC group. Subplot (B) shows that from an intramodular perspective, compared to the HC group, there were fewer connections within the SMN network. * Indicates that the results can survive Bonferroni correction (*P<*0.05).

## 4. Discussion

The current study explored the PC features in all ASD subjects, in different age groups, and in different subtype groups. The current study unravels the brain features of ASD and provides new insights into understanding the neural mechanism of ASD.

### 4.1. ASD subjects show poorer network segregation in the DMN than healthy controls

The current study found a higher mean PC in the ASD group than in the HC group. Further analyses revealed that the increased PC was caused by lower intranetwork connectivity and higher internetwork connectivity in ASD patients than in HCs. Specifically, for intramodular connectivity, there were fewer connections within the DMN and SMN network in the ASD group than in the control group. For intermodular connectivity, the ASD group showed significantly increased FC between the DMN and the FPN (or CON).

We considered the modular structure of brain networks and found transdiagnostic changes in ASD patients ^33^. Based on previously reported network-level studies, an increasing participation coefficient represents the characteristics of network integration, and a decreasing participation coefficient represents the characteristics of network segregation ^34^. High PC reflects the characteristics of network integration and indicates an enhanced role in coordinating messaging in brain networks, which may reflect pathological adaptations ^35^. Thus, in the current study, the increase suggests that ASD subjects show less developed network segregation.

For intranetwork connectivity, further analysis shows that the reduction in the number of connections within the network results in a lower degree of modular segregation ^33^. Previous studies have consistently reported that DMN modules become increasingly separated ^36-39^. In psychiatric disorders, schizophrenia exhibits lower intra-SMN connectivity than major depressive disorder, suggesting a greater reduction in local functional connectivity in the sensorimotor cortex in schizophrenia ^40^. In the current study, the reduction of the intranetwork connections in ASD indicated lower modular specialization in the DMN and lower local functional connectivity in the sensorimotor network in the SMN. For internetwork connectivity, a meta-analysis article demonstrated the existence of hyperconnectivity between the DMN and FPN networks, which is a common feature of different psychiatric disorders ^41^. The anterior DMN network showed an increased correlation (hyperconnectivity) with the cingulo-opercular network ^5^. The change in the structure of modules in ASD is mainly due to the excessive number of intermodule connections involving higher-order and primary modules, which indicates dedifferentiation of the network organization.

Taking all the intra- and internetwork results into consideration, we can conclude that ASD subjects show decreased intranetwork connections and increased internetwork connections, which result in higher PC results. The results suggest that ASD subjects show poorer network segregation than healthy controls, which made them show impaired cognitive or behavioural functions versus healthy controls, and this feature may be responsible for their disorders. These changes hindered the ASD brain from performing higher-level functions, integrating higher-level information, and differentiating and processing specific information.

### 4.2. Poor network segregation in the DMN was found in primary school-aged and adolescent ASD subjects but not in adult subjects

**With respect to** different age groups, the current study found a higher mean PC in the ASD group than in the HC group in primary school children and adolescents. Further analyses revealed that the increased PC in the primary school group was caused by lower intranetwork connectivities in ASD, and the increased PC in adolescents was caused by lower intranetwork connectivities and higher internetwork connectivities in ASD. In addition, we observed that the differential relationship between modularity in ASD among primary school-aged participants and adolescents was mainly due to changes in the SMN intranetwork. In other words, the SMN may have driven the increase in modularity of ASD over time. This may also be the main reason for the increase in modularity from primary school age to adolescence ^14^.

In addition, no group difference was observed in adult ASD patients and HCs. This feature also provides an explanation of the features in adult ASD. Studies have shown that the short-range connections between brain regions gradually decrease and the long-range connections gradually increase with the development of PC results ^42-45^. Specifically, decreased functional connectivity of the DMN is observed in adults aged 36-86 years during normal ageing ^46^. The DMN becomes increasingly segregated as it ages through strengthening of the intranetwork connections ^44^. These changes allow the brain to perform higher-level functions, to integrate higher-level information and to differentiate and process specific information. As a result, there may be fewer internetwork connections and more intranetwork connections, which is consistent with our results.

All of these results are consistent with the results for all subjects and are consistent with the modularity of ASD rising from children to adolescents and then falling from adolescents to adults ^14^. The current results provide an explanation of the neural features underlying the developmental features in adult ASD.

### 4.3. The network segregation features in subtypes of ASD

Regarding the subtypes of ASD, the current study found a higher mean PC in the DMN in the ASD group than in the HC group in Type I and Type III. Further analyses revealed that the increased PC in Type I was caused by lower intranetwork connectivity and higher internetwork connectivity in ASD. Although there was no significant difference in PC for type II patients, ASD patients showed a significantly lower value in the SMN intranetwork than HCs. These results first supported the results for all subjects and also suggest that although there are differences in different types of ASD, there are commonalities in neural features among the different subtypes.

In Asperger’s syndrome (type II ASD), participants show some specific features of types I and III. Studies revealed that these participants scored lower on image sensory and orientation tasks related to the interpretation of tactile and proprioceptive information and lower with respect to postural exercises indicative of perceptual-motor impairment, which itself is associated with sensory processing problems ^47^. One study found that Asperger’s syndrome and high-functioning autism patients showed significant motor impairment compared to controls. Meanwhile, there was no significant difference between the HFA and Asperger’s syndrome groups ^48^. All of these articles demonstrated the specific features of Type II, which might be the reason for the difference in subtypes. In the current study, we observed that the differential relationship between modularity and Type II was mainly due to changes in the SMN intranetwork. This may also be the main reason for the absence of network integration features in Type II ^14^. Future studies should focus on the subtype features of ASD and explore the similarities and differences among different types of ASD.

### 4.5. Limitations

Several limitations of this study need to be noted. First, PC measurements may be affected by the size of the module ^1^. When focusing on the absolute results of PC analysis, more advanced PC calculation methods should be considered. Second, there is no uniformity in the classification of subtypes in either DSM-IV or DSM-5, and a more refined classification should be required for future analyses. Finally, the study should be more specific to each brain region for intranetwork analysis and should consider the issue of directionality for internetwork analysis.

## Conclusions

The current study found that the disruption of DMN network topology in ASD included consistent partial contributions from module-specific levels of integration and specialization changes, which provides new insights into the contribution of module-specific changes in ASD to network dysfunction. In terms of age, the ASD group was characterized by a quadratic pattern, and the modularity of ASD rose from children to adolescents and then fell from adolescents to adults. Among the subtypes of ASD, we observed not only commonality between the different types (Type I and Type III have the same biomarkers) but also differences (no network integration features on the DMN for Type II). These findings contribute to our understanding of the underlying neurological basis of ASD and have important implications for the development of effective interventions to treat ASD.

## Data Availability

The data stored at our lab-based network attachment system: http://QuickConnect.cn/others. ID:guests; PIN

## Author contribution

Bo Yang analyzed the data, prepared the figures, tables and wrote the methods and results sections. Min Wang, Weiran Zhou, and Shuaiyu Chen contributed to research idea and data analyses techniques; Xiuqin Wang and Lixia Yuan contributes to data-preprocessing and multi-site counterbalancing; Marc Potenza and Guang-Heng Dong designed this research and edited the manuscript. All authors contributed to and approved the final manuscript.

## Acknowledgements

This research was supported by The Cultivation Project of Province-levelled Preponderant Characteristic Discipline of Hangzhou Normal University (20JYXK008), and Zhejiang Provincial Natural Science Foundation (LY20C090005). The funding agencies did not contribute to the experimental design or conclusions, and the views presented in the manuscript are those of the authors and may not reflect those of the funding agencies. This article has been posted on the preprint server MedRxiv.

## References

1. Hill EL, Frith U. Understanding autism: insights from mind and brain. Philos Trans R Soc Lond B Biol Sci. 2003;358(1430):281–289.

2. Paakki JJ, Rahko J, Long X, et al. Alterations in regional homogeneity of resting-state brain activity in autism spectrum disorders. Brain Res. 2010;1321:169–179.

3. Weng SJ, Wiggins JL, Peltier SJ, et al. Alterations of resting state functional connectivity in the default network in adolescents with autism spectrum disorders. Brain Res. 2010;1313:202–214.

4. Yerys BEG, E. M. Abrams, D. N. Satterthwaite, T. D. Weinblatt, R. Jankowski, K. F. Strang, J. Kenworthy, L. Gaillard, W. D. Vaidya C. J. Default mode network segregation and social deficits in autism spectrum disorder: Evidence from non-medicated children. Neuroimage Clin. 2015;9:223–232.

5. de Lacy N, Doherty D, King BH, Rachakonda S, Calhoun VD. Disruption to control network function correlates with altered dynamic connectivity in the wider autism spectrum. Neuroimage Clin. 2017;15:513–524.

6. Buckner RL, Andrews-Hanna JR, Schacter DL. The brain’s default network: anatomy, function, and relevance to disease. Ann N Y Acad Sci. 2008;1124:1–38.

7. Uddin LQM, V. Young, C. B. Ryali, S. Chen, T. Khouzam, A. Minshew, N. J. Hardan A. Y. Multivariate searchlight classification of structural magnetic resonance imaging in children and adolescents with autism. Biol Psychiatry. 2011;70(9):833–841.

8. Eilam-Stock TX, P. Cao, M. Gu, X. Van Dam, N. T. Anagnostou, E. Kolevzon, A. Soorya, L. Park, Y. Siller, M. He, Y. Hof, P. R. Fan J. Abnormal autonomic and associated brain activities during rest in autism spectrum disorder. Brain. 2014;137(Pt 1):153–171.

9. Lynch CJ, Uddin LQ, Supekar K, Khouzam A, Phillips J, Menon V. Default mode network in childhood autism: posteromedial cortex heterogeneity and relationship with social deficits. Biol Psychiatry. 2013;74(3):212–219.

10. Uddin LQ, Supekar K, Lynch CJ, et al. Salience network-based classification and prediction of symptom severity in children with autism. JAMA Psychiatry. 2013;70(8):869–879.

11. Uddin LQ, Supekar K, Menon V. Reconceptualizing functional brain connectivity in autism from a developmental perspective. Front Hum Neurosci. 2013;7:458.

12. Washington SD, Gordon EM, Brar J, et al. Dysmaturation of the default mode network in autism. Hum Brain Mapp. 2014;35(4):1284–1296.

13. Haghighat H, Mirzarezaee M, Araabi BN, Khadem A. Functional Networks Abnormalities in Autism Spectrum Disorder: Age-Related Hypo and Hyper Connectivity. Brain Topogr. 2021;34(3):306–322.

14. Henry TR, Dichter GS, Gates K. Age and Gender Effects on Intrinsic Connectivity in Autism Using Functional Integration and Segregation. Biol Psychiatry Cogn Neurosci Neuroimaging. 2018;3(5):414–422.

15. Lord C, Petkova E, Hus V, et al. A multisite study of the clinical diagnosis of different autism spectrum disorders. Arch Gen Psychiatry. 2012;69(3):306–313.

16. de Giambattista C, Ventura P, Trerotoli P, Margari M, Palumbi R, Margari L. Subtyping the Autism Spectrum Disorder: Comparison of Children with High Functioning Autism and Asperger Syndrome. J Autism Dev Disord. 2019;49(1):138–150.

17. Besseling RL, R. Michels, B. Heunis, S. de Louw, A. Tijhuis, A. Bergmans, J. Aldenkamp B. Functional network abnormalities consistent with behavioral profile in Autism Spectrum Disorder. Psychiatry Res Neuroimaging. 2018;275:43–48.

18. McPartland J, Volkmar FR. Autism and related disorders. Handb Clin Neurol. 2012;106:407–418.

19. Servaas MNR, H. Renken, R. J. Wichers, M. Bastiaansen, J. A. Figueroa, C. A. Geugies, H. Mocking, R. J. Geerligs, L. Marsman, J. C. Aleman, A. Schene, A. H. Schoevers, R. A. Ruhe H. G. Associations Between Daily Affective Instability and Connectomics in Functional Subnetworks in Remitted Patients with Recurrent Major Depressive Disorder. Neuropsychopharmacology. 2017;42(13):2583–2592.

20. Yang XL, J. Meng, Y. Xia, M. Cui, Z. Wu, X. Hu, X. Zhang, W. Gong, G. Gong, Q. Sweeney, J. A. He, Y. Network analysis reveals disrupted functional brain circuitry in drug-naive social anxiety disorder. Neuroimage. 2019;190:213–223.

21. Pedersen MO, A. Walz, J. M. Zalesky, A. Jackson, G. D. Spontaneous brain network activity: Analysis of its temporal complexity. Netw Neurosci. 2017;1(2):100–115.

22. Floris DL, Filho JOA, Lai MC, et al. Towards robust and replicable sex differences in the intrinsic brain function of autism. Mol Autism. 2021;12(1):19.

23. Van Dijk KR, Hedden T, Venkataraman A, Evans KC, Lazar SW, Buckner RL. Intrinsic functional connectivity as a tool for human connectomics: theory, properties, and optimization. J Neurophysiol. 2010;103(1):297–321.

24. Yan CG, Chen X, Li L, et al. Reduced default mode network functional connectivity in patients with recurrent major depressive disorder. Proc Natl Acad Sci U S A. 2019;116(18):9078–9083.

25. Jenkinson M, Bannister P, Brady M, Smith S. Improved Optimization for the Robust and Accurate Linear Registration and Motion Correction of Brain Images. NeuroImage. 2002;17(2):825–841.

26. Wang C, Hu Y, Weng J, Chen F, Liu H. Modular segregation of task-dependent brain networks contributes to the development of executive function in children. Neuroimage. 2020;206:116334.

27. Dosenbach NU, Nardos B, Cohen AL, et al. Prediction of individual brain maturity using fMRI. Science. 2010;329(5997):1358–1361.

28. Redcay E, Moran JM, Mavros PL, Tager-Flusberg H, Gabrieli JD, Whitfield-Gabrieli S. Intrinsic functional network organization in high-functioning adolescents with autism spectrum disorder. Front Hum Neurosci. 2013;7:573.

29. Wang J, Wang X, Xia M, Liao X, Evans A, He Y. GRETNA: a graph theoretical network analysis toolbox for imaging connectomics. Front Hum Neurosci. 2015;9:386.

30. Amaral RGLsAN. Functional cartography of complex metabolic networks. Nature. 2005;433(7028):892–895.

31. Nielson DM, Pereira F, Zheng CY, et al. Detecting and harmonizing scanner differences in the ABCD study—annual release. 2018.

32. Gordon EM, Laumann TO, Gilmore AW, et al. Precision Functional Mapping of Individual Human Brains. Neuron. 2017;95(4):791–807 e797.

33. Ma Q, Tang Y, Wang F, et al. Transdiagnostic Dysfunctions in Brain Modules Across Patients with Schizophrenia, Bipolar Disorder, and Major Depressive Disorder: A Connectome-Based Study. Schizophr Bull. 2020;46(3):699–712.

34. Lopez KC, Kandala S, Marek S, Barch DM. Development of Network Topology and Functional Connectivity of the Prefrontal Cortex. Cereb Cortex. 2020;30(4):2489–2505.

35. Gong Q, He Y. Depression, neuroimaging and connectomics: a selective overview. Biol Psychiatry. 2015;77(3):223–235.

36. Kohno M, Morales AM, Dennis LE, McCready H, Hoffman WF, Korthuis PT. Effects of Naltrexone on Large-Scale Network Interactions in Methamphetamine Use Disorder. Front Psychiatry. 2019;10:603.

37. Liang X, He Y, Salmeron BJ, Gu H, Stein EA, Yang Y. Interactions between the salience and default-mode networks are disrupted in cocaine addiction. J Neurosci. 2015;35(21):8081–8090.

38. Stopyra MA, Simon JJ, Skunde M, et al. Altered functional connectivity in binge eating disorder and bulimia nervosa: A resting-state fMRI study. Brain Behav. 2019;9(2):e01207.

39. Wall MB, Pope R, Freeman TP, et al. Dissociable effects of cannabis with and without cannabidiol on the human brain’s resting-state functional connectivity. J Psychopharmacol. 2019;33(7):822–830.

40. Wei Y, Chang M, Womer FY, et al. Local functional connectivity alterations in schizophrenia, bipolar disorder, and major depressive disorder. J Affect Disord. 2018;236:266–273.

41. Sha Z, Wager TD, Mechelli A, He Y. Common Dysfunction of Large-Scale Neurocognitive Networks Across Psychiatric Disorders. Biol Psychiatry. 2019;85(5):379–388.

42. Iidaka T. Resting state functional magnetic resonance imaging and neural network classified autism and control. Cortex. 2015;63:55–67.

43. Fair DA, Dosenbach NU, Church JA, et al. Development of distinct control networks through segregation and integration. Proc Natl Acad Sci U S A. 2007;104(33):13507–13512.

44. Gu S, Satterthwaite TD, Medaglia JD, et al. Emergence of system roles in normative neurodevelopment. Proc Natl Acad Sci U S A. 2015;112(44):13681–13686.

45. Supekar K, Musen M, Menon V. Development of large-scale functional brain networks in children. PLoS Biol. 2009;7(7):e1000157.

46. Yasuhiro Funakoshi MH, Hideki Otsuka_2_, Kenji MOri, Hiromichi Ito, and Takashi Iwanaga. Default mode network abnormalities in children with autism spectrum disorder detected by resting-state functional magnetic resonance imaging. 2016.

47. Siaperas P, Ring HA, McAllister CJ, et al. Atypical movement performance and sensory integration in Asperger’s syndrome. J Autism Dev Disord. 2012;42(5):718–725.

48. Jansiewicz EM, Goldberg MC, Newschaffer CJ, Denckla MB, Landa R, Mostofsky SH. Motor signs distinguish children with high functioning autism and Asperger’s syndrome from controls. J Autism Dev Disord. 2006;36(5):613–621.

